# Dietary diversity practice and associated factors among pregnant women attending ANC in Kolfe Keranyo sub city health centers, Addis Ababa, Ethiopia

**DOI:** 10.1101/2020.04.27.20081596

**Authors:** Walelgn Tefera, Tsegahun Worku Brhanie, Mamo Dereje

**Affiliations:** Kolfe Keranyo sub city health centers, Addis Ababa, Ethiopia; Bahir Dar University, Ethiopia

**Keywords:** DDS, Pregnant women, Income, ANC, Education and Nutrition information

## Abstract

**Background:** Adequate and healthy diet during pregnancy is essential for the health of both mother and newborn. Dietary diversity is a proxy indicator of maternal nutrient adequacy. However, little is documented on dietary diversity among pregnant women.

**Objectives:** This study was designed to assess the dietary diversity practice and associated factors among pregnant women attending ANC in health centers of the coffee keranyo sub city, Addis Ababa, Ethiopia, 2018

**Methods:** Institution based cross-sectional study was conducted on 406 randomly selected pregnant women attending ANC in health centers of the coffee keranyo sub city, Addis Ababa from March 2-April 2/ 2018. Data were collected by using interviewer and 24 H dietary recall method. Data had entered and analyzed using SPSS version 21. Multiple logistic regression was run to assess factors associated with the dependent variable at P<0.05.

**Result:** The mean DDS was 5.45± 1.83. About 60.9% of pregnant women had good dietary diversity practice. Pregnant women learned collage and above had more dietary diversity practice than the illiterate one [AOR=2.26., 95% CI: (1.066, 4.808)]. Pregnant women with monthly income more than 5,000 ETB had more dietary diversity than income less than 2,000 ETB [AOR=2.33, 95% CI: (1.234, 4.416)]. Pregnant women at second ANC visit had more dietary diversity than at first visit [AOR=2.42, 95% CI: (1.183, 4.952)]. Having nutrition information during pregnancy increases 2 times dietary diversity practice than none informed ones [AOR=2.10, 95% CI: (1.294, 3.422)].

**Conclusion and Recommendation:** The mean DDS among the pregnant mothers was 5.45. 60.9% of pregnant women had good dietary diversity score and 39.1 % had poor dietary diversity. Mothers education, monthly income, second and third ANC visit and nutrition information had a positive significant with pregnant mothers’ dietary diversity (P<0.05. Early initiation of ANC visit and incorporation of nutrition education in each visit should be practice. Health extension workers should provide nutritional education to every pregnant woman.

## 1. Introduction

### 1.1 Background

Dietary diversity refers to an increase in the variety of foods across and within food groups capable of ensuring adequate intake of essential nutrients that can promote good health, and physical and mental development(1).

There is no any single food which contains all the required nutrients for optimal health. (2) And a diversified diet is associated with a good nutritional status (3). A variety of foods in the diet has therefore been considered important in ensuring adequate intake of essential nutrients and in realizing an optimal nutritional status (4). Dietary diversity has been shown to be strongly and positively correlated with nutrient adequacy in low-income countries. Nutrient adequacy is one important element of diet quality. Thus, the Minimum Dietary Diversity of Women can be used as a proxy for this aspect of diet quality. It can be used to monitor and evaluate programs that seek to improve diet quality in resource-constrained settings (5). Lack of dietary diversity is a challenge for rural communities in developing countries. Their diets are by default defined on starchy staples with inadequate animal products, fresh fruits and vegetables (6).

During pregnancy a woman needs good nutritional status for a healthy outcome. Women who have a poor nutritional status at conception are at higher risk of disease and death; their health depends greatly on the availability of food, and they may be unable to cope with their increased nutrient needs during pregnancy in situations of food insecurity. Infections such as malaria, HIV and infestation with gastrointestinal parasites can exacerbate such women’s under nutrition (7).

During pregnancy all women need more food, a varied diet, and micronutrient supplements. When energy and other nutrient intake do not increase, the body’s own reserves are used, leaving a pregnant woman weakened. Energy needs increase in the second and particularly the third trimester of pregnancy. Inadequate weight gain during pregnancy often results in low birth weight, which increases an infant’s risk of dying Pregnant women also require more protein, iron, iodine, vitamin A, folate, and other nutrients. Deficiencies of certain nutrients are associated with maternal complications and death, fetal and newborn death, birth defects, and decreased physical and mental potential of the child. Pregnant women need to consume extra vitamins and minerals, increase their calorie intake, and avoid certain foods such as chemicals to optimize the growth and development of their baby (8).

Poor maternal nutrition, both before and during pregnancy, is associated with adverse pregnancy outcomes including intrauterine growth restriction, which greatly increases the risk of neonatal deaths, low birth weight (LBW), preterm birth (PTB), and stunting. Thus, improving the dietary pattern and nutritional status both before and during pregnancy can play a major role in preventing anemia, intrauterine growth restriction, and the associated short- and long-term adverse effects. This was in line with the current emphasis on the first 1000 days of life as a window of opportunity to promote healthy child growth (9). Maternal micronutrient malnutrition is a widespread nutrition challenge faced by women living in resource-poor settings, the consequences of which affect not only the health and survival of women, but also that of their children, notably through intrauterine growth retardation (10).

One of the main factors responsible for this type of malnutrition is the poor quality of women’s diets as they lack dietary diversity. There is ample evidence from developing countries that dietary diversity is indeed strongly associated with nutrient adequacy (11).

In resource-poor environments across the globe, low-quality, monotonous diets were the norm. When grain or tuber-based staple foods dominate and diets lack vegetables, fruits and animal-source foods, the risk for a range of micronutrient deficiencies was high. Women of reproductive age (15–49 years old) are particularly vulnerable because of their greater micronutrient needs (12). In Ethiopia Deficiencies in micronutrients such as folate, iron and zinc and vitamins A, B6, B12, C, E and riboflavin are highly prevalent and may occur concurrently among pregnant women. Multiple micronutrient supplementations in pregnant women may be a promising strategy for reducing adverse pregnancy outcomes through improved maternal nutritional and immune status (9). Improving the quality of women’s diet is the best way to stop the intergenerational cycle of malnutrition (13).

The ‘minimum dietary diversity-women’ is a global indicator recently endorsed to monitor nutrition-sensitive actions and programs aimed at improving the diet of women of reproductive age by UN agencies, FAO and FANTA in 2015. According to the MDD-W, women who have consumed at least 5 of the 10 possible food groups over a 24-hour recall period are classified as having minimally adequate diet diversity. The Food and Agriculture Organization (FAO) and the United States Agency of International Development (USAID) both recommend the use of the MDD-W when a categorical indicator of individual dietary diversity for women is needed. These organizations also recommend the use of the WDDS, although it has no universally recommended cut-off value to indicate scale of dietary diversity and can only be used as a continuous variable (14).

Simple, rapid, and useful proxy measures and indicators like the FAO’s scoring system for measuring Women’s Dietary Diversity Score (WDDS) have been shown to be valid proxy indicators for various nutritional monitoring activities. Therefore, efforts to enhance maternal awareness about nutrition in general, and promoting dietary diversity (≥ 5 food groups) to increase animal sources foods (ASF), fruits and vegetables in particular, could help improve maternal nutrition and prevent the associated adverse outcomes (15).

### 1.2 Statement of the problem

Maternal and child under nutrition, is the leading global developmental challenge affecting nearly half of the world’s population and responsible for the death of 3.5 million mothers and children annually (16). Now, developing countries are burdened with the ‘triple burden of malnutrition’ encompasses the three dimensions of under nutrition (wasting, stunting & underweight), micronutrient deficiencies and over nutrition. Food security policies should focus not only on calorie intake but also consumption of a diversified diet (17).

Maternal malnutrition has been strongly linked to functional consequences like increased risk of adverse pregnancy outcomes, poor infant survival and risk of chronic diseases at later stages of life. Studies showed that nutrition during pregnancy was the single most important factor predicting maternal anemia, preterm birth, intrauterine growth restriction and reproductive loss through still births (9). It is also the major determinant factor in the risk of giving low birth weight infants, the burden of both maternal and child under nutrition along with the adverse outcomes is skewed to few countries in low and middle income regions in Africa and Asia. A major reason for the widespread malnutrition and the associated consequences in Sub-Saharan Africa is the monotonous, plant-based diets that may be inadequate to provide adequate nutrition during pregnancy. Diets in such setting consist mainly of cereal or root staples; with very little intake of animal source proteins, vegetables, and fruits and thus are poor sources of bioavailable mineral and vitamins critical for healthy pregnancy (18).

Malnutrition and poor diets constitute the number-one driver of the global burden of disease. We already know that the annual GDP losses from low weight, poor child growth, and micronutrient deficiencies average 11 percent in Asia and Africa. The impact of El Niño of malnutrition throughout 2015 and into 2016 has been profound, resulting in a monotonous, plant-based diet that may be inadequate to provide adequate nutrition during pregnancy. Diets in such setting consist mainly of cereal or root staples; with very little intake of animal source proteins, vegetables, and fruits and thus are poor sources of bioavailable mineral and vitamins critical for healthy pregnancy (19). Staggering caseload of severe acute malnutrition in eastern and southern Africa and an estimated 60 million people affected overall so far (20).

In Ethiopia, majority of pregnant mothers (59.9%) had a poor dietary practice during pregnancy. They lacked the basic and the essential practice to consume vegetables, fruits, egg and others which are the basic sources of most of the types of vitamins and minerals. Moreover, unhealthy food practice was observed among them (21) and majority (75.2 %) of pregnant women did not take additional meal during pregnancy; about 69.3 % of the pregnant women were skipping one or more of their regular meals (22). The content and quality of the foods consumed by Ethiopian women do not changed when they are pregnant. Some women had decreasing or avoiding consumption of fruits, which they believe will make their fetus “fatter.” During pregnancy there is tendency for some women to decrease their food intake over pre-pregnancy intake for two reasons: During their first trimester of pregnancy, they experience nausea and aversions to certain foods; and During the later stages of pregnancy (late second trimester and the third trimester), some women deliberately decrease their food intake in an effort to have a smaller fetus and an easier delivery. Known in the nutrition literature as “eating down,” this practice has been reported previously in Ethiopia and other countries (23).

In Ethiopia maternal malnutrition results maternal anemia, pre-term birth and low birth weight. A Women dietary diversity score of >=4 food groups during pregnancy was shown to be associated with lower risk of maternal anemia, LBW, and PTB. And women consuming dairy, animal-source foods, fruits, and vegetables including vitamin A–rich ones has good maternal outcome (24). In Ethiopia Large proportion of women (98.3%) relay on monotonous food group and only 10.2%women found in the high dietary diversity score (DDS >6) means eating more than 6 food groups from10. The consumption of essential micronutrients rich foods such as VitA and Iron was very low. The study in west Gojam, shows Pregnant women who eat plant based VitA rich foods = 10.3%, Animal based VitA rich foods = 43.6% (25). There are different factors associated to this poor dietary diversity practice, including household monthly income, educational status of pregnant women (21, 26) and nutrition information of the pregnant women (21,27) and employment status, household assets, land ownership and morbidity(26),family size, food group avoidance and mothers’ occupation (27).

The study in Kenya showed the critical role of education, occupation, monthly income, household assets, land ownership and maternal morbidity status in the attainment of the minimum dietary diversity and ultimately improved nutrient intake among pregnant women (26). In Ethiopia many studies have focused on nutritional status of pregnant women. Consistent evidence about Ethiopian’s pregnant women dietary practice is lacking. Taking into consideration dietary diversity practice that continue to occur throughout pregnancy, it will be informative to investigate and identify the dietary diversity practice that occur among Ethiopian pregnant women and reasons associated with the dietary diversity practice. Therefore, this study aimed to investigate dietary diversity practice and associated factors of women during their pregnancy.

### 1.3 Significance of the Study

The study is important for ministry of health, Addis Ababa Health Bureau, the Kolfe Keranyo sub city and other governmental and nongovernmental organizations working on promotion of maternal health to implement programs aimed at improving dietary diversity among pregnant women. The study is also used for researchers and planners for the secondary source of data. The study has contributed to gain knowledge about dietary diversity and associated factors to individuals.

## 2. Literature review

Malnutrition is one of the most serious health problems affecting children and their mothers in the world. Currently out of7 billion world population of About2 billion people are suffering from micronutrient malnutrition, Nearly 800 million people suffer from calorie deficiency. Out of 5 billion adults worldwide, nearly 2 billion are overweight or obese, and one in 12 has type 2 diabetes. Out of 667 million children under age 5 worldwide, 159 million are stunted 50 million are wasted and 41 million are overweight (20). In Ethiopia the problem is huge, according to EDHS 2016 among all under 5 children, 38% are stunted,10% wasted and 24% overweight (28).

Micronutrient malnutrition is among the leading and commonest health problems globally, accounting for 7% of the global disease burden and comes with a global cost of $180 billion each year. One in three people have at least one form of micronutrient deficiency (29). The most common micronutrient deficiency is Iron deficiency anemia (IDA), alone affects more than 2 billion people globally and contributes to over 100,000 maternal and almost 600,000 perinatal deaths each year (30). Contrarily, malnutrition related to micronutrient deficiency is one of the most widespread, yet largely neglected nutritional challenges faced by women living in the developing world (17). The most common cause of micronutrient malnutrition in developing countries is the intake of monotonous cereal based diets that are lacking in diversity. Diets in these countries lack fruits, vegetables and animal source foods, Due to this inadequate nutrient intake among pregnant women, iron deficiency anemia and other micronutrient deficiencies have remained prevalent in developing countries (31). In Ethiopia, even if there is limited evidence on the level of iron deficiency, the prevalence of iron deficiency anemia among pregnant women was around 23% (32).

In Pakistan, study showed dietary diversity is a good proxy indicator for micronutrient adequacy in pregnant women, but the practice was low,22% of pregnant women were overweight, 17% obese, and only 12% of pregnant women were under weight. Similarly, 28.1% were anemic. And 89% of pregnant women had Medium dietary diversity (ate 4–5 food groups from the ten), while only 5% showed low (ate less than 4 food groups from 10) and 6% showed high dietary diversity (ate more than 5 food groups from 10). More than 74% of pregnant women gained less than recommended level of weight gain (33).

In Kenya, Most women (30.3%) had attended their antenatal clinic at least once. Only 22.4% indicated to have attended the antenatal clinic more than three. Interestingly, results revealed that majority (70.1%) of pregnant women were under micronutrient supplementation. Notably, for those under supplementation, majority (98.9%) reported intake of iron and folic supplements. The mean dietary diversity score was 6.84 ± 1.46 SD. Based on the established terciles, most (60.6%) of the respondents were in the high dietary diversity tercile (≥6 food groups). Additionally, 37% were in the medium (4–5 food groups) and 2.4% low dietary diversity tercile (≤3 food groups) (26).

In Ethiopia dietary diversity practice is very low, literatures in different region show poor dietary diversity practice of pregnant women. For example the study in Gondar 59.9%pregnant women had poor dietary practice and 40.1% had good dietary practice (21). Different literature shows that a lack of dietary diversity or consumption 4 or less than 4 food groups from the 10 during pregnancy results in 28.6% to 32.4% maternal anemia, 9.1% low birth weight, 13.6%preterm birth and 4.5% still birth. Evidence shows that involved supplementation with Iron or iron foliate, multiple micronutrients, and, more recently, food supplementation has several advantages in reducing LBW and preterm (9).

The study in West Gojam, shows only 10%of children consumed food group of six and more, Majority of the children (78.6%) consumed cereal based foods (maze, finger millet, teff, wheat, sorghum, barley and oat), Large proportion of women (98.3%) relay on the monotonous food group, only 10.2%women found in the high dietary diversity score (DDS >6). The consumption of essential micronutrient rich foods such as VitA and Iron was very low. Plant based VitA rich foods = 10.3% Animal based VitA rich foods = 43.6% Iron rich foods consumed by children = 16% (25). In Wallega study showed only 33.9% of the pregnant women were found to have good nutritional practices during their pregnancy. Some pregnant mothers in the study (35.8%) had practiced avoiding food during their pregnancy due to religion, culture and thinking that the food items make the baby big and more than half of the respondent had diet frequency of meal one to two per day (27). Study in Mirab Abaya, southern, Ethiopia showed that nearly one third (34.3%) of the household had high dietary diversity while the rest 65.7% had low dietary diversity (34). Study in Bahir Dar town showed only 39.3% of the pregnant women had good dietary practice and the rest 60.7% of pregnant women reported poor dietary practices. (35).

There are different factors associated with dietary diversity practice of pregnant women, includes family monthly income, mother’s educational status and nutrition education.(21,26,35), food availability and access(36), number of trimester, morbidity, personal dislike(food aversion) 18.3% and cultural habits(food taboo)2.6% (27). Sex of head of the household, marital status, monthly expenditure and presence of old age dependency were factors associated with dietary diversity (34). The study in Ghana showed that household wealth index classification and frequent ANC attendance were the only significant independent predictors of maternal dietary diversity (37).

### 2.1 Conceptual framework

There were three dimensions variables which had responsibility for dietary diversity of pregnant women. The first includes variables directly related to pregnant women, including. Age, educational status, marital status, occupation, cultural or personal food avoidance, Religion, nutrition information, Number of pregnancy and trimester The second

## 3. Objectives of the study

### 3.1 General objective

To assess dietary diversity practices and associated factors among pregnant women attending ANC in public health centers of Kolfe Keranyo sub city, from March - April 2018.

### 3.2 Specific objectives: the specific objective of the study were to

✓ To determine the proportion of pregnant women practicing dietary diversity among pregnant women attending ANC in public health centers of Kolfe Keranyo sub city.
✓ To Identify factors associated with dietary diversity among pregnant women attending ANC in public health centers of Kolfe Keranyo sub city.

## 4. Methods and Materials of the research

### 4.1 Description of the study area

The study was conducted in Kolfe Keranyo sub city which is one of the ten sub city of Addis Ababa. Addis Ababa is the capital city of Ethiopia and represented by all ethnic group of Ethiopia. Administratively, the city is divided in to 10 sub cities and 116 woredas. The city has 3.8 million populations and has health service access in less than 2 kilometer for all people. There are 5 hospitals under Ministry of heath, 6 Hospitals under Addis Ababa Health Bureau, 2 Hospitals under Ministry of Defense, 1 Hospital under Police Force and around 40 Private Hospitals. The city also has 95 functional governmental health centers and more than 760 private clinics in the 10 sub cities. Kolfe Keranyo sub city is found in the western part of Addis Ababa, Ethiopia. Administratively, the sub city is divided in to 15 districts. There are 1 hospital under Ministry of health and 3 Private Hospitals. The sub city also has 11 functional governmental health centers and more than 176 private clinics in the sub cities (38). Based on Kolfe Keranyo sub city health office, the total population of the sub city in 2017 is estimated to be around 547,940 and has a growth rate of 3.8% and population density of 4,165 persons per square kilometer. The sex ratio shows 48.5% are males and 51.5% are females. It had an estimated number of pregnancy 12, 603 (38).

### 4.2 Study Design

Institutional based cross-sectional design was employed to assess dietary diversity among pregnant women attending ANC in public health centers of Kolfe Keranyo sub city, Addis Ababa.

### 4.3 Source and Target Population

#### 4.3.1 Source population

The source populations were all pregnant women attending ANC in public health centers of Kolfe Keranyo sub city.

#### 4.3.2 Target Population

All Pregnant women who visit the randomly selected health facilities during March to April, 2018 were considered as the study population

### 4.4 Inclusion and exclusion criteria

#### 4.4.1 Inclusion criteria

All pregnant women attending ANC in the study facilities and being resident of Kolfe Keranyo sub city for at least 6 months were included.

#### 4.4.2 Exclusion criteria

All pregnant women attending ANC in the study facilities and currently live out of Kolfe Keranyo sub city were excluded.

### 4.5 Study Variables

**Dependent Variable:** Dietary diversity practice of pregnant women with

**Independent Variables:** age, educational status, health status, religion, occupation, number of ANC visit, nutrition information, parity, trimester, family size, Income level and food item avoidance.

### 4.6 Operational Definitions

**Dietary diversity**: Refers to the number of food groups consumed by pregnant women over a 24-hour period.

**Low or poor dietary diversity score**: When pregnant women consumed less than five food groups among the 10 groups within 24 h before the survey.

**High or good dietary diversity score:** When pregnant women consumed five or more food groups among the 10 groups within 24 h before the survey.

**Minimum Dietary Diversity of pregnant women**: Dietary diversity score of pregnant women receiving at least five food groups out of ten.

**Dietary adequacy**: taking any amount of each food group is considered as adequate.

**Poor household**: households with monthly income less than 2,000 ETB.

**Medium household**: households with monthly income from 2,000–5,000 ETB

**Rich household**; households with monthly income more than 5,000 ETB

### 4.7 Sample Size Determination

The required sample size was determined using the following assumption

**Assumption 1:** Of the proportion of dietary diversity practice, based on the previous study in Gondar, the prevalence of dietary practice is p=40.1% (21). Where: n = required sample sizes, 10% = non - respondent rate Zα/2 = critical value for normal distribution at 95% confidence interval which equals to 1.96 (Z value at alpha = 0.05), d = an absolute precision (margin of error) = 5%

**For the first objective**:

p1 = Proportion of pregnant women dietary diversity practice (40.1%).

n1= (zα/2)^2^ p (1-p)/d^2^ =(1.9^2^× 0.6 × 0.4)/0.05²=0.921984/0.0025=368.79=369

Dietary diversity practice (P1 = 40.1%) n1 = 369 Adding non-response rate of 10% = (369+369×10%) =406

**For the second objective:**

The sample size required was calculated using two-population proportion formula by assuming; *r= n2/n1=1:1*, Z α/2=the value of the standard normal distribution curve corresponding to level of significance alpha 0.05 = 1.96 and the value of the standard normal distribution curve corresponding to 90% power = 1.21

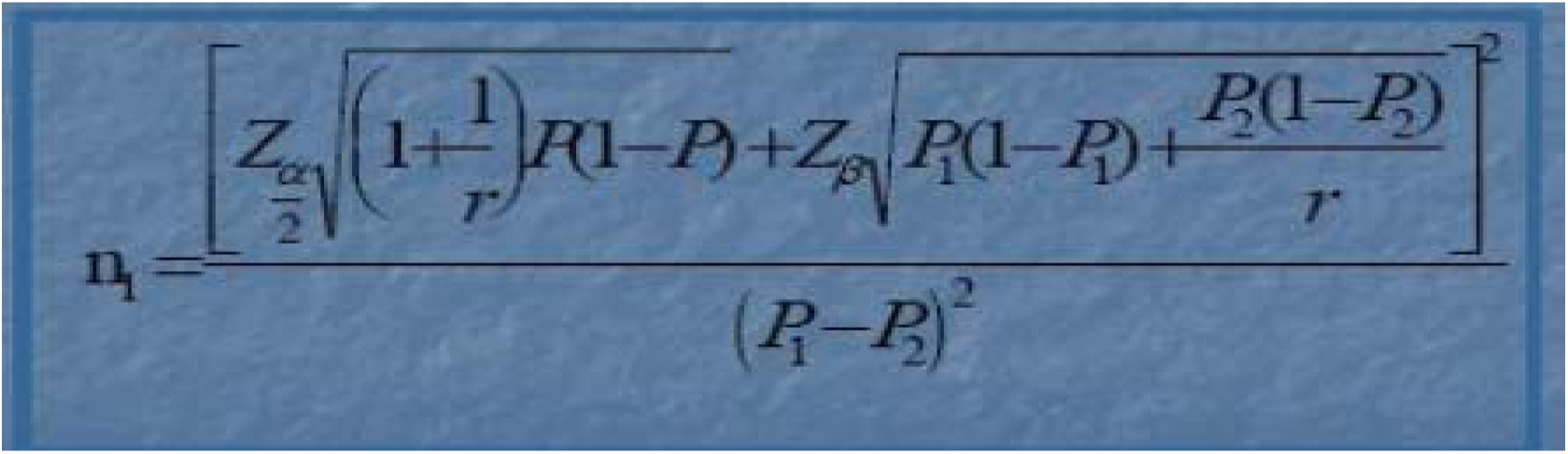

Where: 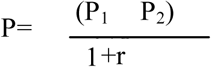
for household income p1 = Prevalence of DDS at household income <1000 birr =25.8%, p2= Prevalence of DDS at household income 1000–2000=51.3%. for educational status p1= Prevalence of DDS among illiterate mothers=23.5%, p2= Prevalence of DDS among mothers learn college and above=52.5%; for nutrition information p1= Prevalence of DDS among mothers who had not nutrition information=20.6%, p2= Prevalence of DDS among mothers who had nutrition information=45.5%;for marital status p1= Prevalence of DDS among unmarried=60%, p2= Prevalence of DDS among married mothers=40%.

**Table 1:** Sampling size determination of associated factors of dietary diversity practice of pregnant women Kolfe Keranyo sub city, Addis Ababa, 2018.

| Variables | DDS P1 (%) | DDS P2 (%) | CI | Power | Allocation ratio | OR | Total sample size | Reference |
| --- | --- | --- | --- | --- | --- | --- | --- | --- |
| Monthly income | 25.8% | 51.3% | 95% | 90% | 1:1 | 2.59 | **163** | Mekonnen Sisay. et al,2014 |
| Education status | 23.5% | 52.5% | 95% | 90% | 1:1 | 2.18 | **121** |  |
| Nutrition information | 20.6% | 45.5% | 95% | 90% | 1:1 | 2.39 | **154** |  |
| Marital status | 60% | 40% | 95% | 90% | 1:1 | 2.62 | **272** | Direslgne Miskir. et al,2016 |

Therefore the first sample size 406 was implemented for pregnant mother who attending antenatal.

### 4.8 Sampling Procedure

A simple random sampling technique was employed to select health centers in the study area. From 11 health centers of sub cities 4 health centers were selected randomly. Then all pregnant women who visit the health center were recorded as a sampling frame in the selected health centers.

Finally, the sample size was proportionally allocated to selected health centers catchment population and estimated pregnancy of Kolfe Keranyo sub city 2018. Respondents were chosen with a systematic random sampling technique with every 2^nd^ pregnant women based on the time of visiting. Kolfe Keranyo sub city has 11 functional health centers namely;- woreda1 (Ayer tena), woreda3(kara), woreda4 (AlemBank), woreda5 (Zenebework), woreda6 (Woira), woreda8 (keranyo),woreda9 (total),woreda11 (philipos),woreda12 (kolfe), woreda13 (lomi meda),and woreda 14 (Birchiko). Catchment population of the sub city has been estimated to 547,940 and estimated pregnancy of 12,603 (38

**Figure 2:**
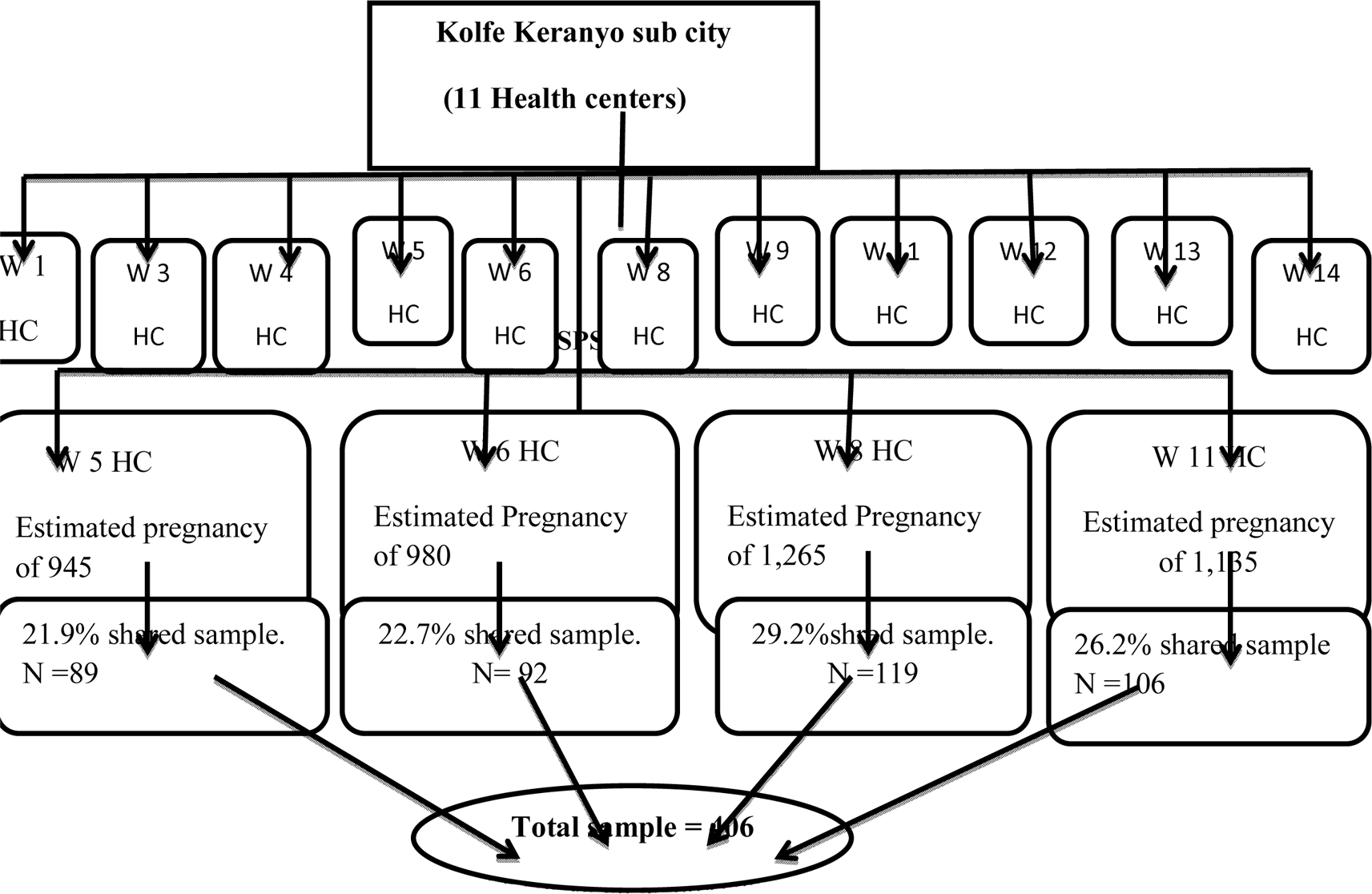
Schematic presentation of sampling procedure

### 4.9 Data Collection Tool and Procedure

Data had collected by exit interview using structured questionnaire and 24 hour recall method adapted from different literatures mainly Food and Agriculture Organization (FAO) Guidelines for measuring household and individual dietary diversity (FAO, 2016) and (Exploring the new indicator Minimum Dietary Diversity-Women Results from Burkina Faso, 2015). A total of ten food groups were considered in this study i.e. Cereals (Grains, white roots and tubers, and plantain), Pulses (beans, peas and lentils), Nuts and seeds, Dairy products, Meat, Poultry, and Fish, Eggs, Dark green leafy vegetables, Other vitamin A-rich fruits and vegetables, Other vegetable and Other fruits (14).

The questionnaire had four main contents: The first includes socio demographic variables of pregnant women, including age, educational status, Religion, occupation, No of pregnancy or parity and trimester. The second group encompasses characteristics of the household, includes family size and household Income level, the third group captured Ante natal care, Health status and nutrition information. The fourth group includes: 24 hour recall feeding practice and dietary diversity. The questionnaire were first prepared in English and then translated to Amharic and translated back to English to observe its consistency. Dietary diversity score (DDS) was collected and calculated as the sum of the number of different food groups consumed by the pregnant women in the 24 H prior to the assessment.

### 4.10 Data quality management

Four clinical nurses were participated in the data collection process. The data collection process was supervised by two senior Public Health professionals and the principal investigators. Both data collectors and supervisors were trained for 1 day about the contents of the questionnaire and on how to collect the data properly in order to minimize errors. The principal investigators and supervisors had made daily supervision during the whole data collection process. The questionnaire was reviewed and checked for completeness, accuracy and consistency by the supervisors and investigators daily and at the end of the whole data collection process.

The questionnaires were pre-tested to check on the length, content, question wording and language. The questionnaire was administered to 5% the sample size, who had attended ANC at non selected (Alem bank) health center. Then error was corrected accordingly.

### 4.11 Ethical clearance

Bahir Dar University Ethical review committee wrote Ethical clearance to the Addis Ababa health bureau to give permission to conduct the research. And Then Ethical clearance was obtained from Ethical review committee of the Addis Ababa health Bureau Then the pregnant women from each selected health centers were informed about the purpose of the study.

**Right of the respondent:** The respondent had the right to refuse in participating prior to the interview, or had the right to withdraw at any time during the interview

**The Risk of the respondent:** Since the procedure was noninvasive and only interview, so there were no personal risk.

**Consent of the respondent**: Verbal consent was obtained prior to data collection from each respondent.

**Privacy and confidentiality:** Privacy and confidentiality of information given by each respondent keep properly and name was not recorded.

**Data security:** The recorded data were coded and kept in a secured place with strict confidentiality.

### 4.12 Data analysis

After all the relevant data was collected, it had entered and then analyzed using SPSS version 21. Using the 10 food groups, dietary diversity categories were formulated namely; low dietary diversity category (<5 food groups) and good or high dietary diversity category (≥5 food groups). The dietary diversity score variable was dichotomized as category 0 for those not meeting the minimum dietary diversity and category 1 for those meeting the minimum diversity.

In the bivariate analysis, the independent variables with a P-value less than 0.2 with the dependent variable were fitted into a multivariable logistic regression model to identify their independent effect on dietary diversity. Independent variables included in the multivariable analysis include mother’s educational status, trimester, and number of ANC visit, nutrition information and household monthly income. The association between the dependent and the independent variables was measured using odds ratio (OR) with 95 % Confidence Interval (C.I). Those variables with p-value of less than 0.05 in the multivariable analysis were considered to be significant as the predictors of minimum dietary diversity.

## 5. Result

### 5.1 Participants’ socio-demographic characteristics

Of the 406 eligible respondents, 402 were willing to respond the questionnaire, which made a response rate of 99%. The mean age of the pregnant women was 27.35 ± 7.89 SD with the range of 16–44 years. Most of the women were married (91.5%) and only 26.1% of the respondents learn collage and above and 30.1% of the pregnant women were housewife.

78.4%% pregnant women were not the household heads, about 81(20.2%) of the pregnant women are informal salaried employees.

In regard to family size, the average size was 4 people, with 4.47±1.95 SD. The mean monthly household income was 3686.26 ETB and about 306 (76.1%) of the household earned below 5000 ETB monthly.

**Table 2:**
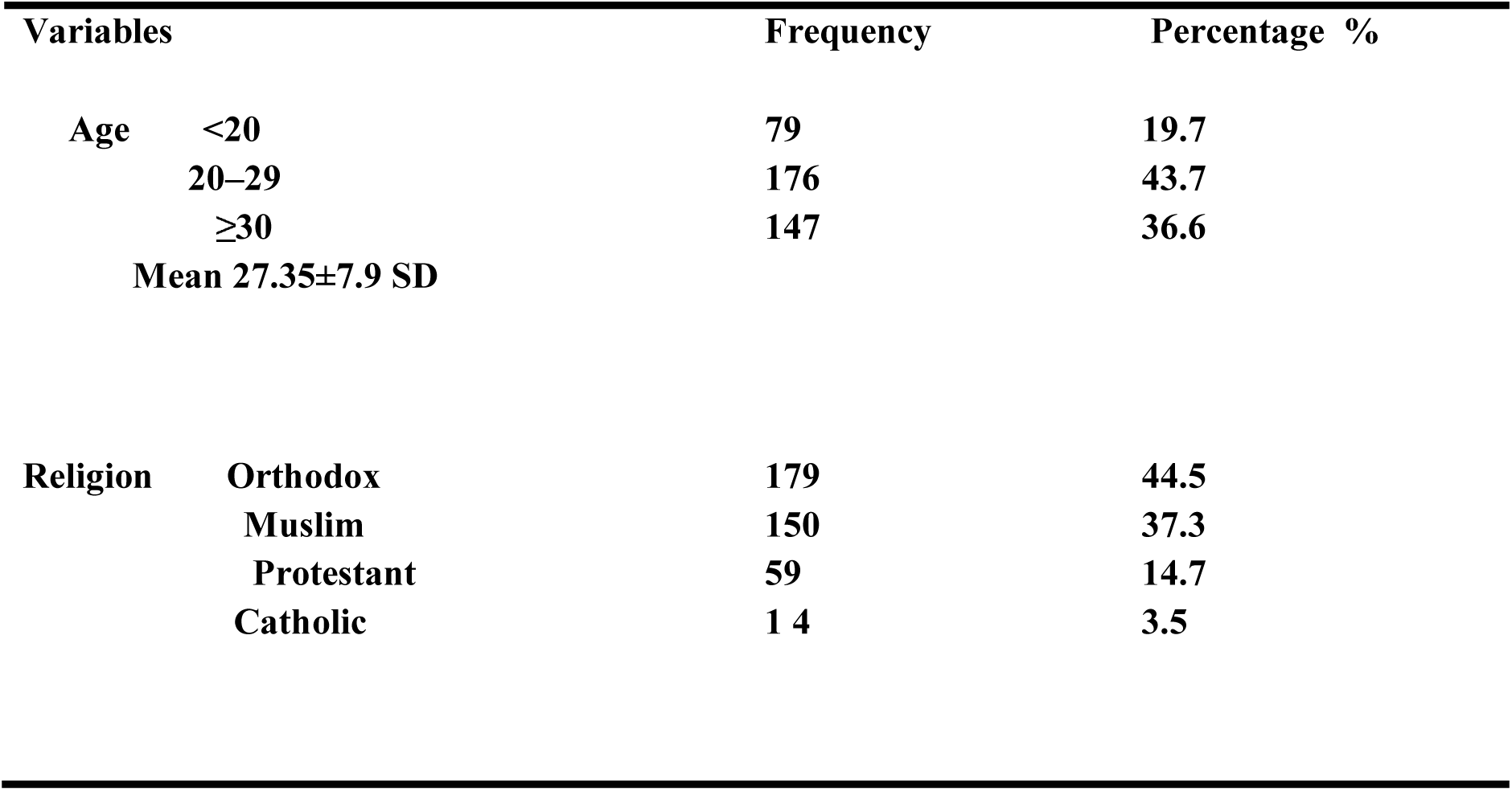
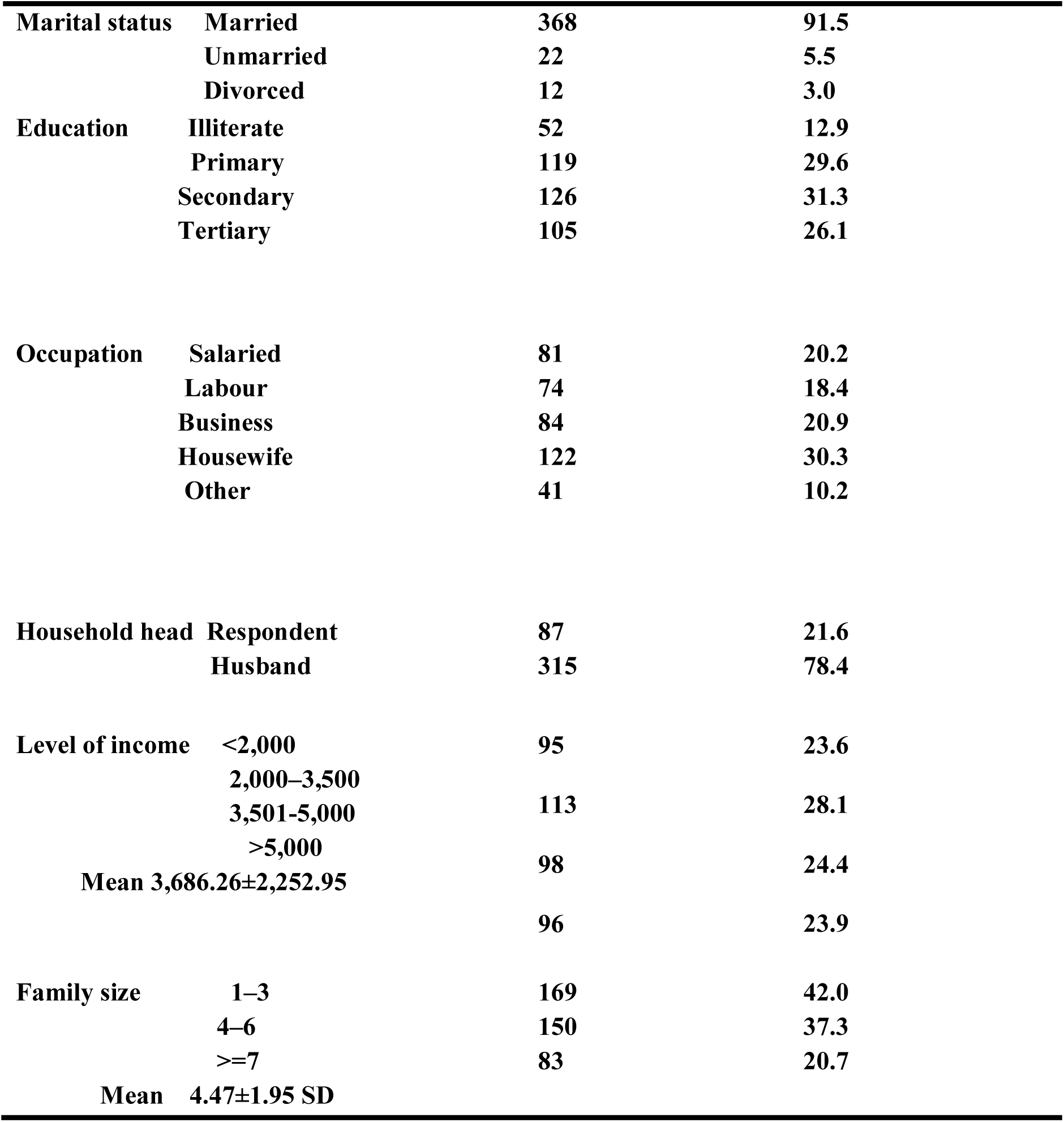
Socioeconomic and Demographic characteristics of Pregnant Women in Kolfe Keranyo sub city, Addis Ababa 2018. (n = 402)

### 5.2 Maternal and nutrition characteristics

The mean parity of the study respondents was 3 children with the range 1–7. More than half (51.2%) of the pregnant women were in their second trimester and about 165 (41.0%) of the respondents were at second ANC visit during the interview. About 190 (47.3%) of having nutrition information about the importance of dietary diversity during pregnancy, And the ANC visit was the main source of information for 63% of mothers.

Notably, in regard to maternal morbidity, the study found that 117 (29.1%) of the pregnant women reported some form of illness/disorder in the preceding two weeks prior to the day of the interview. The most commonly reported disorders were dyspepsia/heartburn by 37% of the study participants. Due to cultural or personal dislike 126 (31.3%) of pregnant women had avoided at least one food item from their meal. Of these egg 46%, dark green leafy vegetables 27% and milk and milk products 17%.

**Table 3:**
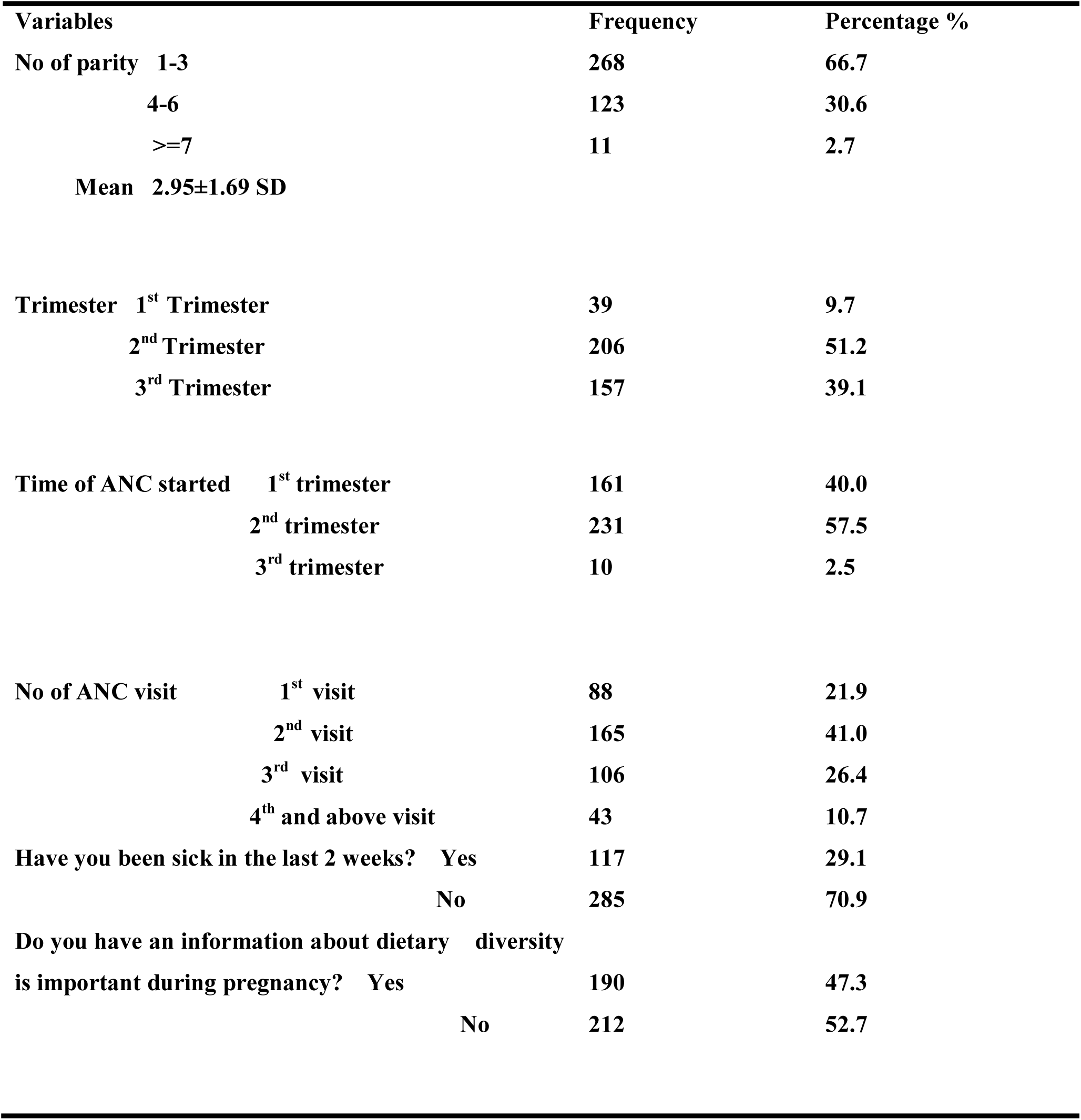
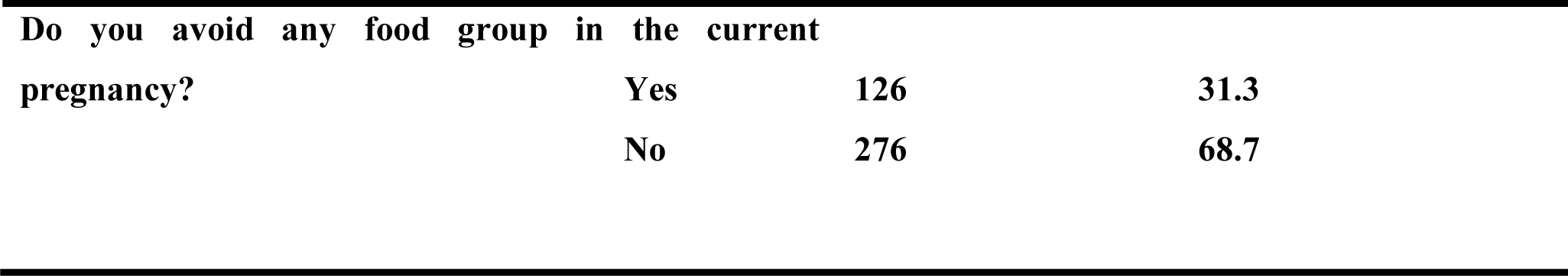
Maternal and nutrition characteristics of pregnant women in Kolfe Keranyo sub city, Addis Ababa 2018. (n = 402)

### 5.3 24 hours dietary diversity practices of pregnant mothers

#### 5.3.1 Dietary diversity score of the pregnant women

Out of the 10 food groups, the study found the mean DDS was 5.45± 1.83 SD with scores ranging from 3 to10 food groups. Based on the categories developed, Nearly 2/3^rd^ (60.9%) of respondents had a good dietary diversity score (≥5 food groups) and about 39.1% of the participants were in the low diversity category (<5 food groups).

The most commonly eaten foods were cereals (100%) and pulses were the second most eaten food group 63.9%. Notably, the egg was minimally consumed 36(8.9%).

**Table 4:** Respondents by food group, pregnant women in kolfe keranyo sub city, Addis Ababa 2018. (n = 402)

| **Food group** | **n = 402** | **Percentage** |
| --- | --- | --- |
| Cereals (Grains, white roots and tubers, and plantain) | 402 | 100 |
| Pulses (beans, peas and lentils) | 257 | 63.9 |
| Nuts and seeds | 132 | 32.8 |
| Dairy products | 149 | 37.0 |
| Meat, poultry, and fish | 189 | 47.0 |
| Eggs | 36 | 8.9 |
| Dark green leafy vegetables | 173 | 43.0 |
| Other vitamin A-rich fruits and vegetables | 129 | 32.0 |
| Another vegetable | 253 | 62.9 |
| Other fruits | 88 | 21.8 |

**Figure 2:**
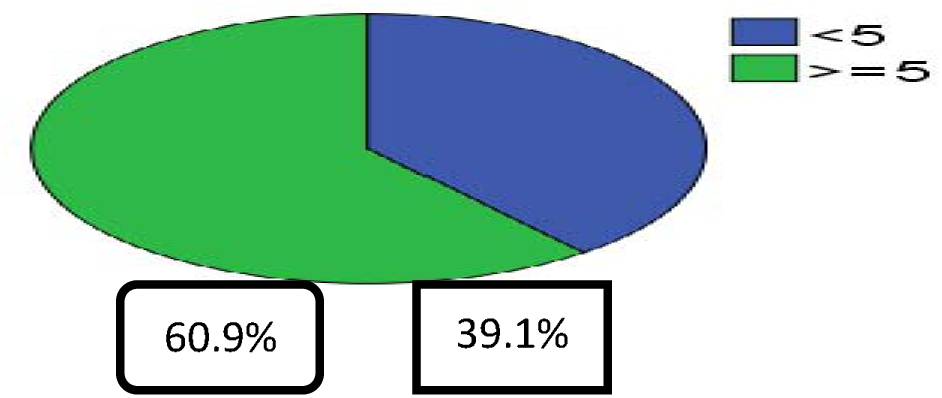
Minimum dietary diversity practice of pregnant women,Kolfe Keranyo subcity Addis Ababa 2018.(n=402).

### 5.4 Factors associated with dietary diversity practice of pregnant women

Bivariate analysis showed that there was an association between dietary diversity practices of pregnant mothers and mother’s educational status, trimester, number of ANC visit, nutrition information and household monthly income, whereas, age, marital status, religion, household head, mothers occupation, parity, family size, time of ANC start, avoidance of food items and sickness had no association with dietary diversity practices of mothers. The result table 5 of, multivariate analysis showed that there was statistically significant (p <0.05) assocciation between educational status, family monthly income, number of ANC visit and nutrition information dietary diversity practices of pregnant mothers.

**Table 5;.** Multivariable logistic regression analyses showed factors associated with dietary diversity practice among pregnant women in Kolfe Keranyo sub city, Addis Ababa 2018. (n = 402)

| Variables | Practice |  | COR with 95%CI | P value | AOR with 95%CI |
| --- | --- | --- | --- | --- | --- |
|  | ≥5 (High) | <5 (Low) |  |  |  |
| Education |  |  |  |  |  |
| Illiterate | 22 (5.5) | 30 (7.5) | 1.00 |  | 1.00 |
| Primary | 72(17.9) | 47 (11.7) | 2.09 (1.08 , 4.05) | 0.033 | **2.16 (1.06-4.39)** |
| Secondary | 79 (19.7) | 47 (11.7) | 2.29 (1.19 , 4.43) | 0.034 | **2.13 (1.06-4.30)** |
| Collage and above | 72 (17.9) | 33 (8.2) | 2.98 (1.50 , 5.92) | 0.033 | **2.26 (1.07-4.81)** |
| Income |  |  |  |  |  |
| <2000 | 45 (11.2) | 50 (12.4) | 1.002.108 |  | 1.00 |
| 2000-3500 | 74 (18.4) | 39 (9.7) | (1.2053.688) | 0.056 | 1.78 (0.98-3.22) |
| 3501-5000 | 59 (14.7) | 39 (9.7) | 1.681 (0.950-2.975) | 0.208 | 1.48(0.80-2.75) |
| >5000 | 67 (16.7) | 29 (7.2) | 2.567 (1.418-4.646) | 0.009 | **2.33 (1.23-4.42)** |
| Trimester |  |  |  |  |  |
| 1<sup>st</sup> trimester | 19 (4.7) | 20 (5) | 1.00 |  | 1.00 |
| 2nd trimester | 112 (27.9) | 94 (23.4) | 1.25 (0.63 , 2.49) | 0.201 | 0.56 (0.23-1.36) |
| 3rd trimester | 114 (28.4) | 43 (10.7) | 2.79 (1.36 , 5.73) | 0.674 | 0.80 (0.29-2.23) |
| Number of ANC attendance |  |  |  |  |  |
| 1st visit | 35 (8.7) | 53 (13.2) | 1.00 |  | 1.00 |
| 2nd visit | 104 (25.9) | 61 (15.2) | 2.58 (1.52 , 4.39) | 0.016 | **2.42 (1.18-4.95)** |
| 3rd visit | 73 (18.2) | 33 (8.2) | 3.35 (1.85 , 6.06) | 0.046 | **2.30 (1.01-5.21)** |
| 4th and above | 33 (8.2) | 10 (2.5) | 4.99 (2.19 , 11.42) | 0.058 | 2.87 (0.96-8.51) |
| Having nutrition information |  |  |  |  |  |
| Yes | 142 (35.3) | 48 (11.9) | 3.13 (2.05 , 4.78) | 0.003 | **2.10 (1.29-3.42)** |
| No | 103 (25.6) | 109 (27.1) | 1.00 |  | 1.00 |

Pregnant women learned collage and above had 2.3 times, good dietary diversity practice than the illiterate mothers [AOR=2.264., 95% CI: (1.066, 4.808)]. Pregnant women with household income more than 5,000 00 ETB had dietary diversity practice almost 2.3 times than those households having a household income less than 2000 per month [AOR=2.334, 95% CI: (1.234, 4.416)]. Pregnant women who were at second ANC visit had 2.4 times more attaining minimum dietary diversity than who were at first visit [AOR=2.420, 95% CI: (1.183, 4.952)].

There was also statistical association between nutrition information and minimum dietary diversity practices of mothers during pregnancy. Having nutrition information during pregnancy increases 2 times dietary diversity practice of pregnant women than none informed ones [AOR=2.104, 95% CI: (1.294, 3.422)].

## 6. Discussion

In the present study, a mean DDS of 5.45± 1.83 SD was reported in table 4. 60.9% of pregnant women had good dietary diversity practice and 39.1% pregnant women did not receive a minimum dietary diversity, which is better than similar study conducted in Kenya, reported mean DDS 4.6 (36). But this mean DDS is lower as compare to literatures conducted in different countries, Another study in Kenya showed mean DDS of 6.84 ± 1.46 SD and 60.6% of pregnant women received minimum dietary diversity was reported (26). Similar study done in Pakistan and South Africa among pregnant women, where a mean DDS of 6.17 ± 0.99 (39) and 6.70 ± 2.22 SD (40) was recorded, respectively. This result was also greater than study conducted in Bahir Dar town, North West Ethiopia; showed that only 39.3% of the pregnant women had good dietary practice and the rest 60.7% of pregnant women reported poor dietary practices (35). Another study done in Gondar, Ethiopia near to this study area reported 40.1% as good dietary practice of pregnant women (21). Similarly, same study conducted in Gojam, North West, Ethiopia showed, that only 47% of women were received ≥4 food group from 10 food groups (25). This difference is may be due to different cultural, educational and economic back ground and different study period for seasonal availability of different food groups.

In this study table 4, around 100% and 63.9 % of the pregnant mothers had consumed cereals (grains, white roots and tubers, and plantain) and Pulses (beans, peas and lentils), respectively in the previous 24 hrs. Conversely, egg and other fruit food group were least consumed food groups (8.9% and 21.8%, respectively). Another study in Kenya showed that (99%) pregnant women had eaten cereals, (93%) vegetables and (92%) milk and milk products, but foods of animal origin were minimally consumed only 4% (26). Similar finding are observed in south Achefer, West gojam, Ethiopia, large proportion of women (98.3%) relay on monotonous food group (25).

This study also showed that 31.3% of pregnant women avoid at least one food group from their meal. The reason to avoidance was either personal dislike or cultural beliefs that makes the fetus so big or causes maternal diarrhea. Similar study in Wollega, south west Ethiopia reported 35.8% (27). Study in Bahir Dar town, North West Ethiopia; indicated that 203(33%) pregnant women avoid certain foods, of which 74.4% avoids food due to religious reason (35).

In regard to the association between dietary diversity and the selected maternal demographic factors (age, marital status, trimester of the pregnancy, occupation, and household size), this study did not find any statistically significant association. Similar, findings were also documented in Kenya, except occupation those who reported being employed (salaried) had the highest odds (2.29 times) of attaining minimum dietary diversity as compared to the non-employed (26) and in Gondar, Ethiopia reported that no association (21). But, study in Mirab Abaya, Southern, Ethiopia, reported that age and marital status had statistically significant association with pregnant women dietary diversity (34).

Multivariable analysis showed that there was statistically significant association (p<0.05) between maternal educational status, family monthly income, second and third ANC visit and nutrition information with dietary diversity practices of pregnant mothers.

Pregnant women with household income more than 5,000 ETB had dietary diversity practice almost 2.3 times than those households having household income less than 2000 per month. And in Kenya those households with higher income had better chances of having diversified diets, those with an income of between Kenyan shilling (KSh) 20, 000 and KSh30, 000 were 2.01 times more likely to attain the minimum diversity as compared to those who had an income of less than 10, 000 (26). This association is the same to study in Gondar, Ethiopia, dietary practice during pregnancy among mothers who had monthly income of 1000–2000 was 2 times higher than those <1000 monthly income [AOR=2.18, 95% CI: (1.39, 3.39)] (21). Slightly strong association also observed in Bahir Dar, Ethiopia, pregnant women earn more than 2000 ETB monthly were 3.1 times more likely to have good dietary practice than those earning less than 1000 ETB [AOR = 3.12,95% CI, 91.743, 5.586)] (35)

Pregnant women learned college and above had 2.3 times more dietary diversity practice than the illiterate mothers. Other related study also showed association between women education and dietary diversity practice, in Kenya, study observed that those who had tertiary [AOR 2.93; 95% CI 1.40, 8.63) and secondary education [AOR 2.78; 95% CI (1.06, 5.32)] had greater odds of achieving the minimum dietary diversity as compared to those who had never attended school (26). Which is supported by the study conducted in Gondar identified that educational status have strong statistical association with dietary practices of mothers during pregnancy [AOR=2.59, 95% CI: (1.38, 4.85)] (21).

There is also statistical association between nutrition information and dietary practices of mothers during pregnancy. Having nutrition information during pregnancy increases 2 times dietary diversity practice of pregnant women than none informed ones. Similar study showed strong association in Wollega, Western, Ethiopia, reported that Women who had information about nutrition during pregnancy had 6.3 more likely good nutrition practice than women who had no information during pregnancy [AOR= 6.26, 95% CI: (3.49–11.25)] (27) and in Gondar, Ethiopia, also there was statistical association between nutrition information and dietary practices during pregnancy [AOR=2.39, CI: 1.44–3.97] (21). In Bahir Dar, Ethiopia, pregnant women who had nutrition information were 3.17 times more likely to have good dietary practices than their counterparts [AOR = 3.17, 95%CI, (1.76, 5.67)] (35).

Pregnant women who were at second ANC visit [AOR=2.420, 95% CI: (1.183, 4.952)] and third ANC visit [AOR 2.300, 95% CI: (1.014–5.213)] had more attaining minimum dietary diversity than who were at first visit. In Ghana showed that frequent ANC attendance were the only significant independent predictors of maternal dietary diversity (37). Association may be in three different reasons first; pregnant women in the second and third visit had got enough health investigation, screening and nutritional advice during first visit. Secondly, pregnant women at first visit had not information about dietary diversity and they might have hyperemesis which causes loss of appetite. Thirdly, pregnant women at fourth visit assumed that eating protein diet at term leads the fetus big and not such important.

## 7. Conclusion and Recommendation

### 7.1 Conclusion

The mean DDS among the pregnant mothers was 5.45 and 60.9% pregnant women had good dietary diversity practice.

All pregnant women, mostly rely on Cereals (Grains, white roots and tubers, and plantain), however, egg, other fruit and other vitamin A-rich fruits and vegetables were minimally eaten.

Factors like maternal educational status, family monthly income, second and third ANC visit and nutrition information were strongly associated with good dietary diversity practices of pregnant mothers.

### 7.2. Recommendation

➢ Health extension workers in house to house visit should focus on providing nutritional education to increase the practices of dietary diversity during pregnancy.
➢ Initiate ANC visit as early as she knows, so as to enhance dietary diversity practice of the women.
➢ Health facilities should incorporate nutrition education with other ANC service in every ANC visit to enhance dietary diversity of pregnant women.
➢ Synergy between health and educational sector so as to increase maternal educational status.
➢ Encourage further research in Ethiopia to determine proportion of national minimum dietary diversity practice of pregnant women.

## Data Availability

Data's wrote in the paper, the data is not available online

## Authors’ Contribution

Walelgn Tefera-wrote the first draft, data collection, analysis and interpretation Tsegahun Worku-correction of the analysis and interpretation, editing and manuscript preparation. Mamo Dereje-Comment and Suggestion. All authors approved the final manuscript.

## Acknowledgements

Thank you for the respondents, data collectors and supervisors for their great role of successful data collection. We would like to express our heartfelt gratitude to Kolfe Keranyo sub city health office administration for their willingness, technical support and encouragement.

## Reference

1. Daniels CM. Dietary diversity as a measure of Women’s diet quality in resource-poor areas: results from metropolitan Cebu, Philippines site. Washington: Food and Nutrition Technical Assistance II Project (FANTA-2);2009.

2. Kennedy, G. Evaluation of dietary diversity scores for assessment of micronutrient intake and food security in developing countries. Wageningen University,2009.

3. Drimie, S., Faber, M., Vearey, J., & Nunez, L. Dietary diversity of formal and informal residents in Johannesburg, South Africa, 2013; 13(1): 911.

4. Ey, C. E., Zalilah, M. S., Ys, C. Y., & Norhasmah, S. Dietary diversity is associated with nutritional status of Orang Asli children in Krau Wildlife Reserve, Pahang. Malaysian Journal of Nutrition, 2012; 18(1):1–13.

5. Arimond, M., Wiesmann, D., Becquey, E., Carriquiry, A., Daniels, M.C., Deitchler, M., Fanou-Fogny, N., Joseph, M.L., Kennedy, G., Martin-Prével, Y. & Torheim, L.E. Simple food group diversity indicators predict micronutrient adequacy of women’s diets in 5 diverse, resource-poor settings. J Nutr. 2010; 140(11): 2059S–69S.

6. Arimond, M. and Ruel, M. (2002), “Summary indicators for infant and child feeding practices: An example from the Ethiopia Demographic and Health Survey 2000”, Food Consumption and Nutrition Division Discussion Paper, Washington, D.C.: International Food Policy Research Institute.

7. World Health Organization, (2012). Nutrition of women in the preconception period, during pregnancy and the breastfeeding period Report by the Secretariat: sixty-fifth world health assembly A65/12; Provisional agenda item 13.3.

8. Maternal Nutrition during Pregnancy and Lactation is a joint publication of LINKAGES: Breastfeeding, LAM, Related Complementary Feeding, and Maternal Nutrition Program and the Child Survival Collaborations and Resources (CORE) Nutrition Working Group. August, 2014

9. Taddese Alemu. Dietary practices of pregnant women and its associations with maternal and perinatal outcomes in rural Central Ethiopia,2016;8:48

10. Allen, L., Multiple micronutrients in pregnancy and lactation: an overview, American Journal of Clinical Nutrition, 2005, 81(5), 1206S-12S.

11. Ruel, M.T. Operationalising dietary diversity: a review of measurement issues and research priorities, Journal of Nutrition, 2003; 133 (11 Suppl. 2): 3911–26.

12. Torheim, L.E., et al., Women in resource-poor settings are at risk of inadequate intakes of multiple micronutrients, Journal of Nutrition, 2010,

13. VArimond, M., Wiesmann, D., Becquey, E., Carriquiry, A., Daniels, M., Deitchler, M., Fanou, N., Ferguson, E., Joseph, M., Kennedy, G., Martin-Prével, Y., Torheim, L.E., Dietary diversity as a measure of the micronutrient adequacy of women’s diets in resource-poor areas: summary of results from five sites, Food and Nutrition Technical Assistance FANTA-2 Bridge and Food Health International, FHI 360, Washington, D.C., 2011.ol. 140, pp. 2051S-8S.

14. Custodio E., Kayitakire F. and Thomas A.C.(2015).; Exploring the new indicator minimum dietary diversity-women. Results from Burkina Faso; EUR 27717; doi:10.2788/86023; FAO. Guidelines for measuring household and individual dietary diversity. Rome: Food and Agriculture Organization of the United Nations; 2016.

15. FAO Summary Report, Meeting to reach consensus on a global dietary diversity indicator for women, Washington, D.C., 2014;7:15–16.

16. Abdel-Razeq, S. & Buhimschi, I. Interpretation of Amniotic Fluid White Blood Cell Count in “Bloody Tap” Amniocenteses in Women With Symptoms of Preterm Labor. Obstet Gynecol, 201;,116;344–354.

17. Bain, L.E. 2013. Malnutrition in Sub-Saharan Africa: burden, causes and prospects. The Pan African medical journal, 15, p.120. World Health Organisation, 2013. Trends in Maternal Mortality: 1990 to 2013.

18. WHO, UNICEF, UNFPA, The World Bank and the United Nations Population Division. Trends in maternal mortality:1990 to 2015 Geneva:World health organization,2015.

19. Imamura, F. et al. Dietary quality among men and women in 187 countries in 1990 and 2010: The Lancet Global Health 2015;3(3):132–142.

20. Global nutrition report, 2016

21. Mekonnen Sisay., Endalamaw Memgesha. Dietary Practice and Associated Factors among Pregnant Women in Gondar Town North West, Ethiopia, 2014. International Journal of Nutrition and Food Sciences, 2015;4(6),: 707–712

22. Kuche Desalegn, Singh Pragya, Moges Debebe. Dietary practices and associated factors among pregnant women in Wondo genet district, southern Ethiopia: A cross-sectional study, 2015;4(5):270–275.

23. USAID, ENGINE and save the children;maternal diet and nutrition practices and their determinants engine: Empowering New Generations to Improve Nutrition and Economic opportunities A project supported by the Feed the Future and Global Health Initiatives A report on formative research findings and recommendations for social and behavior change communication programming in the Amhara, Oromia, SNNP and Tigray regions of Ethiopia; April 2014

24. Taddese A. Melaku U. and Kaleab B; Dietary diversity during pregnancy is associated with reduced risk of maternal anemia, preterm delivery, and low birth weight in a prospective cohort study in rural Ethiopia. AJCN. First published ahead of print May 11, 2016;10:115.116.

25. Abel Ahmed. 2014.; Assessment of Dietary diversity among pregnant and lactating women and 6 to 23 months age children, in rural areas of western Gojjam, Amhara Region

26. Willy K, Judith K, Peter C. Dietary diversity, nutrient intake and nutritional status among pregnant women in Laikipia county, Kenya, 2016; 6(4):378–385.

27. Gemeda Daba, et al. Assessment of Nutritional Practices of Pregnant Mothers on Maternal Nutrition and Associated Factors in Guto Gida Woreda, East Wollega Zone, Ethiopia, 2013; 2(3): 105–113

28. Central Statistical Agency (CSA) [Ethiopia] and ICF. 2016. Ethiopia Demographic and Health Survey 2016: Key Indicators Report. Addis Ababa, Ethiopia, and Rockville, Maryland, USA. CSA and ICF.

29. FAO, 2013. FAO Statistical Yearbook 2013: World food and agriculture. FAO Statistical Yearbook 2013: World food and agriculture, pp.1–307.

30. Kassebaum, N.J. et al., 2014. A systematic analysis of global anemia burden from 1990 to 2010;123(5):615–624.

31. Daniels, Melissa C. (2009). Dietary Diversity as a Measure of the Micronutrient Adequacy of Women’s Diets: Results from Metropolitan Cebu, Philippines Site. Washington, DC: Food and Nutrition Technical Assistance II Project, FHI 360.

32. Alemu Taddese & Umeta M. Reproductive and Obstetric Factors Are Key Predictors of Maternal Anemia during Pregnancy in Ethiopia: Evidence from Demographic and Health Survey (2011). 2015: pp.1–10.

33. Fatima A, Inayat T, Shahzad A. assessment of dietary diversity and nutritional status of pregnant women in islamabad, pakistan, 2014; 26(4):48.

34. Direslgne Misiker. Begosew Misker. Gistane Ayele.; House hold dietary diversity and associated factors in Mirab Abaya wereda Southern Ethiopia: Diversity and Equality in Health and Care, 2016; 13(4): 293–296

35. Amanuel N. and Tona Z.: Dietary practices and associated factors during pregnancy in northwestern Ethiopia, 2018; 18: 183

36. Lydiah M. Waswa. Improving dietary diversity and nutritional health of women and children under two years through increased utilization of local agrobiodiversity and enhanced nutrition knowledge in Kenya, 2016.

37. Mahama S. Maternal Dietary Diversity and Infant Outcome of Pregnant Women in Northern Ghana. International Journal of Child Health and Nutrition, 2012;1: 148–156

38. Kolfe Keranyo sub city health office report,2017

39. Ali F, Thaver I, Khan SA. Assessment of dietary diversity and nutritional status of pregnant women in Islamabad, Pakistan, 2014;26(4):506–9.

40. Acham H, Oldewage-Theron W, Egal AA. Dietary diversity, micronutrient intake and their variation among black women in informal settlements in South Africa: a cross-sectional study. Intern J Nutr Metab. 2012;4:24–39.

